# White adipose tissue inflammation is not attenuated by short-term calorie restriction in obese humans

**DOI:** 10.1101/19005934

**Authors:** Julia Sbierski-Kind, Knut Mai, Jonas Kath, Anke Jurisch, Mathias Streitz, Leon Kuchenbecker, Karsten Jürchott, Leonard Spranger, Reiner Jumpertz von Schwartzenberg, Anne-Marie Decker, Ulrike Krüger, Hans-Dieter Volk, Joachim Spranger

## Abstract

Obesity is a growing global health problem due to its association with chronic low-grade inflammation contributing to metabolic complications. Multiple studies indicate that white adipose tissue (WAT) inflammation can promote type 2 diabetes. However, the function and regulation of both innate and adaptive immune cells in human WAT under conditions of obesity and calorie restriction (CR) is not fully understood yet. Using a randomized interventional design, we investigated postmenopausal obese women who either underwent CR for three months followed by a 4 weeks phase of weight maintenance or had to maintain a stable weight over the whole study period. A comprehensive immune phenotyping protocol was conducted using validated multiparameter flow cytometry analysis in blood and subcutaneous WAT (SAT) (n=21). The T cell receptor repertoire was analyzed by next generation sequencing (n=20) and cytokine levels were determined in SAT (n=22). Metabolic parameters were determined by hyperinsulinemic-euglycemic clamp and then correlated to immune cell subsets. We found that insulin resistance (IR) correlates significantly with a shift towards the memory T cell compartment in SAT. Among various T cell subsets, predominantly CD8^+^ effector memory T cells were associated with obesity-related IR. Interestingly, T cell receptor analysis revealed a diverse repertoire in SAT arguing against an antigen-driven intra-SAT expansion of effector memory T cells. Surprisingly, neither inflammatory cytokine levels nor leucocyte subpopulations were significantly altered upon CR. Our findings demonstrate the accumulation of effector memory T cells in obese SAT contributing to chronic inflammation. The long-standing effect of obesity-induced changes in SAT was demonstrated by preserved immune cell composition after short-term CR induced weight loss.

## Introduction

Obesity is a complex disorder involving an excessive amount of body fat. It is associated with reduced quality of life and with the leading causes of death worldwide, including cardiovascular diseases, stroke, diabetes and some types of cancer (*1-4*). The prevalence of obesity has risen dramatically over the past decades. Today, 2.1 billion people – nearly 30% of the world’s population – are either obese or overweight (*5*). Obesity-associated IR promotes type 2 diabetes (T2D), when exhausted islet β cells fail to compensate for the increased need for insulin to maintain glucose homeostasis (*6*). Low-grade inflammation of hypertrophic WAT plays an etiologic role in the development of IR (*7-9*). Specifically, it has been found that excessive energy intake, accompanied by chronically elevated levels of blood glucose, triglycerides and free fatty acids, seems to alter components of the immune system (*10, 11*). These immunological changes include altered levels of cells of the innate immune system, particularly macrophages (*12, 13*). It has been established that the polarization into pro-inflammatory “M1” or alternatively activated “M2” macrophages influences adipose tissue homeostasis with M1 macrophages promoting inflammation while M2 macrophages help to maintain adipose tissue homeostasis, including insulin sensitivity in lean adipose tissue (*14*). Mast cells, neutrophils and dendritic cells are shown to exacerbate IR (*15-17*), whereas eosinophils and innate lymphoid cells seem to be protective (*16*). Whereas previous research mainly focused on innate immunity, adaptive immune cells (T- and B cells) have emerged as important regulators of glucose homeostasis. They exert inflammatory (including CD8^+^ T cells, Th1, Th17, and B cells) or regulatory influences (including regulatory T (Treg) cells, Th2 cells) and can both exacerbate or protect against IR (*18-20*). In particular, it has been found that the helper T cell composition in peripheral blood correlates significantly with the homeostasis model of assessment for insulin resistance index (HOMA-IR) (*21*) and other measures of adiposity, inflammation and glucose intolerance (*22, 23*). T cells have also been identified in human WAT (*24-27*), but studies elucidating their relationship to local and systemic inflammation and IR and the impact of CR on them are limited. Whereas obesity has been clearly linked to IR and T2D, CR is known to reduce the incidence of heart and kidney disease, hypertension and neurological disorders, to increase sensitivity to insulin, and to be associated with increased longevity (*28, 29*). It is unclear if obesity-related immune cell alterations could be reversed by CR. Studies in the murine spleen, lung, liver and lymph nodes recently found that long-term CR preserves a naïve T-cell phenotype and an immature NK cell phenotype as mice age (*30*). Moreover, is has been reported that both exercise and CR modulate innate and adaptive immunity as well as cytokine levels in inguinal adipose tissue and the spleen in mice (*31, 32*). On the other side, long-term CR is thought to decrease the production of peripheral B lymphocytes and to impair immune function (*33*). Conversely, observations in human studies supported the notion that 6-month CR enhances T-cell–mediated immune response in peripheral blood (*34*). In another multi-center, randomized clinical trial it has been reported that long-term moderate CR reduced circulating inflammatory markers (*35*). Contrastingly, previous work has provided evidence that long-term CR is not associated with T cell immunosenescent markers (*36*). Altogether, there is limited and partially controversial information on the effect of CR on the adaptive immune system in human adipose tissues. To better characterize leucocytes, including T cells, that accumulate in SAT with obesity, to investigate the impact of CR and to further assess potential mechanisms of weight regain after CR, we conducted a sub-study in 21 participants of a randomized controlled clinical trial with obese participants. Here, we compared leucocyte subsets in peripheral blood and SAT, as well as insulin and glucose metabolism and determined the levels of cytokine secretion in SAT directly after the CR induced weight loss and after a period of maintaining the reduced weight. In addition, the TCR-repertoire of intra-SAT T cells was analyzed by next generation sequencing. On the basis of previous animal experiments (*18, 19*), we hypothesized that cytokine levels as well as inflammatory leucocyte cell numbers would correlate with metabolic measures. Moreover, we expected that CR would impact inflammation in obese probands directly after weight loss, demonstrated by altered numbers of leucocytes and inflammatory cytokines.

## Materials and Methods

### Study Design and Adipose Tissue Sampling

In overweight or obese (body mass index (BMI) > 27 kg/m^2^) probands, whole blood and SAT biopsies were sampled before (M0), after weight loss induced by a 12-week low-calorie diet (M3) and after having maintained weight stable over a 4 week period following an isocaloric diet (M4). SAT biopsies were collected by percutaneous needle biopsy of periumbilical fat depots using a 15-gauge needle through skin and anesthetized with 1% lidocaine, as described before (*37*). Blood was obtained at approx. 8 a.m. at time points 0, 3 and 4 months, collected into vacutainers (Thermo Fisher Scientific) containing EDTA (Sigma-Aldrich) for anticoagulation and stored at 4°C for maximum 4 hours. Anthropometry was performed by trained staff under standardized conditions. Results were compared to an obese control group instructed to maintain their weight during the course of the study. Furthermore, blood test results at baseline were compared to a lean control group. Fig. 1A illustrates an overview of the study.

**Figure 1.**
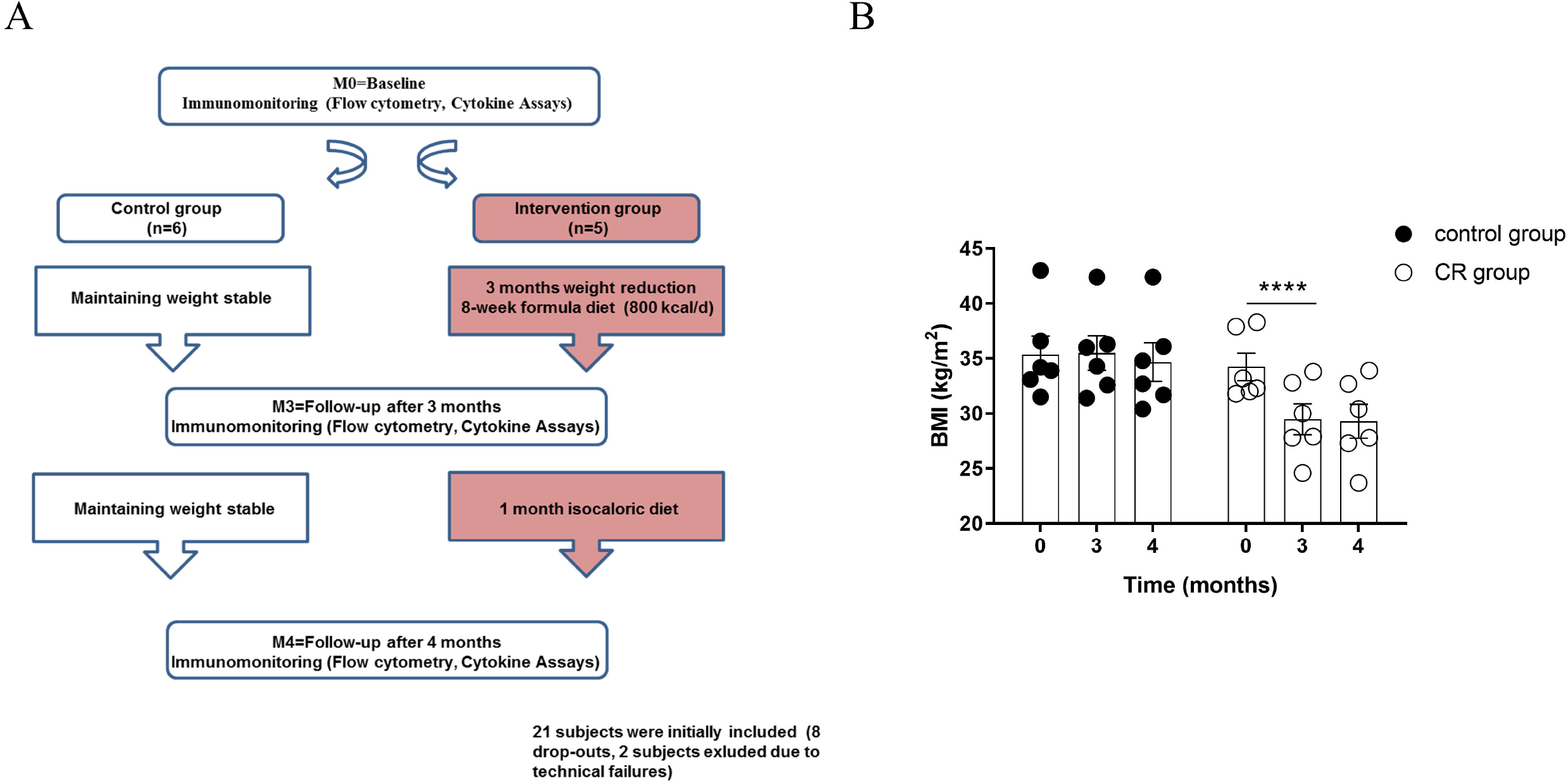
**(A)** Study design: Obese postmenopausal women randomized into two groups (CR group and control group) underwent baseline measurements and follow-up measurements including medical examination and metabolic phenotyping. Between M0 and M3 (after 12 weeks), participants in the CR group lost more than 10% of their body weight following CR. Weight loss was induced by a low-caloric formula diet (800 kcal/day) in the first 8 weeks. Between M3 and M4 (after additional 4 weeks), participants in the CR group followed an isocaloric diet without having a negative energy balance. Participants in the control group were instructed not to change their dietary habits. **(B)** BMI changes of participants of the sub-study C are shown and are representative for the other sub-studies. A repeated measurement 2way ANOVA followed by SIDAK correction testing for group x time interactions indicated significant weight loss in the CR group (**** *P* < 0.0001) between M0 and M3. Participants in the CR group were able to maintain reduced weight until M4 (no significant difference in BMI between M3 and M4). n = 6 per group.

### Participants

The study was performed as a sub-study of a larger study focusing on muscle mass regulation “Effects of negative energy balance on muscle mass regulation” (registered at https://clinicaltrials.gov, NCT01105143) at the Department of Endocrinology of the Charité University Medicine Berlin, Germany. The study was conducted in compliance with the International Conference on Harmonization Guidelines for Good Clinical Practice and the Declaration of Helsinki. All subjects provided written informed consent before participating in this study. The add-on protocol of the study was approved by the local Ethics Committee of the Charité - Universitätsmedizin Berlin (EA2/050/10). Inclusion criteria comprised a BMI > 27 kg/m^2^ and postmenopausal status. The presence of concomitant immunological illness, history of severe untreated medical, neurological, and psychiatric diseases which may interfere with the planned interventions, such as instable coronary heart disease, kidney and liver disease, systemic infections, endocrinological disorders, and hypertension (systolic blood pressure > 180 mm Hg, diastolic blood pressure > 110 mm Hg) were excluded by medical history assessment in obese and lean subjects. Exclusion criteria were also changing dieting or smoking habits significantly in the last two months including a weight loss of 5 kg or more. Cytokine levels of SAT lysates were analyzed in a subgroup of 22 probands before (M0), directly after the calorie restriction period (M3) and after the weight maintenance period (M4) **(sub-study A)**. Blood and SAT T cell receptor sequencing was performed in a subgroup of 20 probands before the CR period (M0) **(sub-Study B)**. Immune cell composition in blood and SAT tissue before and after weight loss was studied in a total of 21 overweight and obese women via flow cytometry (**sub-Study C**). The 21 women were sequentially included into this sub-study, providing additional study consent. Subjects were randomly assigned to the CR group and control group. 8 subjects were not available for all follow-up testing sessions due to personal reasons (half of them were assigned to the CR group). Finally, 11 subjects were available for the per-protocol analysis of sub-Study C (CR group: n=5, control group: n=6). Healthy and lean (BMI < 25 kg/m^2^) age-matched female individuals (n=16) were recruited from staff of Charité - Universitätsmedizin Berlin, Berlin, Germany. The participants gave their written and informed consent prior to participating in the study. The study was approved by the institutional review board (IRB: Ethikkommision der Charité). Characteristics of the subjects analyzed for immune cell composition tested via flow cytometry before and after weight loss are shown in Table 1. Subjects from both CR group and control group were comparable with regard to age, BMI, waist circumference and leucocyte numbers (all *P* > 0.05). Characteristics of subjects analyzed for SAT cytokine secretion are shown in Table 1. Characteristics analyzed for T cell receptor sequencing are also shown in Table 1. Subjects from all groups were comparable with regard to age.

**Table 1.**
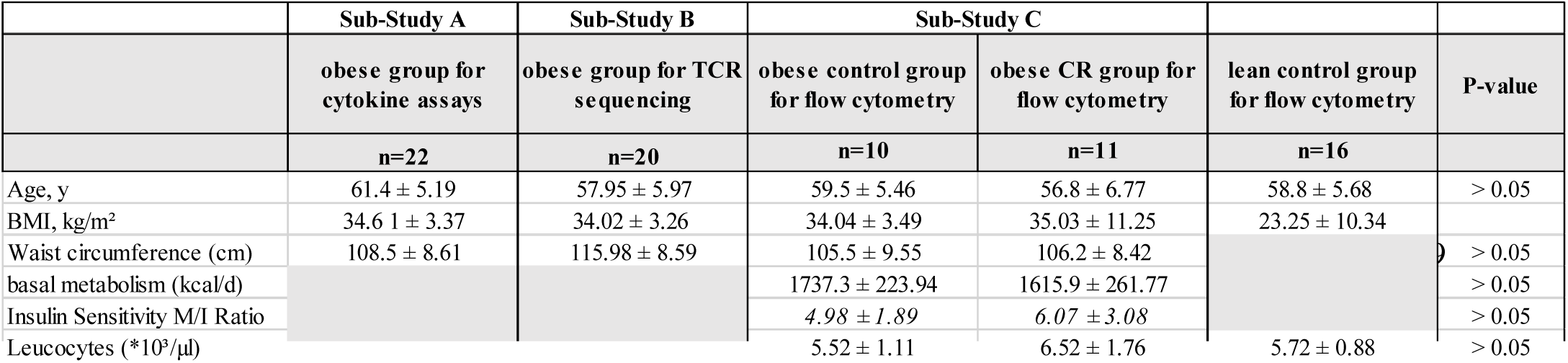
Metabolic and anthropometric parameters of the randomized participants before CR-induced weight loss. The results are presented as mean□±□standard deviation (SD). One-Way ANOVA was conducted and P-values below 0.05 are defined to be significant.

### Calorie Restriction Intervention

In the intervention group, weight loss was induced by low calorie diets. Furthermore, life-style changes were implemented via weekly counseling sessions by clinical dieticians. In the first eight weeks of the 12-week weight loss phase, weight loss was assisted by a low-calorie formula diet (800 kcal/d) replacing all meals with a formula diet (Optifast 2®, Nestlé HealthCare Nutrition GmbH, Frankfurt/Main, Germany). Participants were advised to consume the formula diet exclusively (daily consumption of five packets with 160 kcal each: 20 g carbohydrates, 14 g proteins, and 3 g fat dissolved in 300 ml water) without consuming any additional food. The eight weeks of formula diet were followed by four weeks in which the formula diet was substituted by a calorie reduced healthy diet to facilitate further weight loss (∼ 1300kcal/day). Meals in this diet consisted of a balanced mix of macronutrients in accordance to the guidelines of the German Society for Nutrition (50-55% carbohydrates, 15-20 % proteins, and 30 % fat). The recommended daily calorie intake during these four weeks was based on the measured resting energy expenditure and adapted with regard to subsequent weight changes. Additionally, participants were encouraged to increase physical activity to 150 min of activity per week at least and step counting devices were handed out to monitor and motivate for physical activity. However, no intervention regarding physical exercise was performed. After the 12-weeks weight loss phase, subjects were advised to maintain reduced weight over additional four weeks (until M4). In this weight maintenance phase, subjects were instructed to follow an isocaloric diet. The isocaloric diet was specified individually and subjects were advised to consume a balanced mix of foods with the following distribution: 50-55 % carbohydrates, 30 % fat, 15-20 % proteins. Figure 1B shows that subjects of the intervention group were able to reduce BMI significantly and to maintain weight thereafter. Subjects in the control group were advised to live a healthy lifestyle but maintain weight during the course of the study.

### Hyperinsulinemic-Euglycemic Clamp Procedure

The hyperinsulinemic-euglycemic clamp was performed as described previously (*38, 39*). Briefly, 40 mIU·m^−2^·min^−1^ human insulin (Actrapid, Novo Nordisk) and a variable infusion of 10% glucose (Serag Wiessner) were used. Capillary glucose concentration was then monitored every five minutes and was maintained between 4.0 and 4.9 mmol/L by varying the glucose infusion rate. Insulin sensitivity was assessed as *M*-value and was calculated by dividing the average glucose infusion rate (milligrams glucose per minute) during the steady state of the clamp by the body weight. The insulin sensitivity index (ISI_Clamp_) was calculated as ratio of glucose metabolized during the steady-state period (*M*-value) to a mean serum insulin concentration (milliunits per liter) in this period during the euglycemic clamp. Blood samples were collected before the clamp and at least two hours after starting the clamp during steady-state conditions. Blood samples were centrifuged, and serum and plasma samples were frozen immediately at −80° C.

### Adipose Tissue Cell Isolation

Cells of the stroma vascular fraction were obtained by collagenase digestion of SAT for 90 minutes at 37° C on a rotation shaker (200 rpm). The homogenate was then filtered and centrifuged twice at 500 g for 10 minutes at 4° C. Stroma vascular fraction cells were resuspended in endotoxin-free PBS (Sigma-Aldrich) supplemented with 2 % fetal calf serum (FCS, PAA) and 0.1 % sodium azide (Sigma-Aldrich) and stored at 4° C until the staining was proceeded.

### Antibody Panel

Fluorochrome-conjugated (FITC, PE, ECD, PC7, APC, APC-A700, APC-A750, PacBlue and KrOrange) anti-human monoclonal antibodies were used to stain 30 different epitopes. All antibodies were obtained from Beckman Coulter, except anti-BDCA-2 and anti-BDCA-3, which were obtained from Miltenyi Biotec, and anti-CCR7, which was obtained from R&D Systems. Six panel matrices for 7- to 9-fluorochrome channels defined and validated by the ONE Study consortium as well as gating strategies were used based on published results (*40*).

### Leukocyte Staining

For staining 100 μL of anticoagulated peripheral blood or 100 µl of SAT stroma vascular fraction cells were stained with surface antibodies for 15 minutes at room temperature in the dark prior to lysis and fixation with VersaLyse + 2.5 % IOTest fixative solution (Beckman Coulter) for 15 minutes in the dark. Intracellular staining was performed with the PerFix-nc Kit (Beckman Coulter) and cells were stained with intracellular antibodies for 1 hour. Lysed cells were washed twice (PBS, and PBS containing 2% FCS and 0.1% sodium azide) prior to acquisition. Cell staining was performed within 4 hours after blood collection as recommended in published results to reduce variability. All samples were measured on 10 color, 3 laser Navios flow cytometers (Beckman Coulter) and acquired data files were analyzed using the Kaluza software, version 1.2 (Beckman Coulter). Cell doublets were excluded using forward scatter time of flight (wide) versus forward scatter integral (area). Leukocytes were defined gating CD45 expression versus side scatter. Absolute counts of the subpopulations were calculated in all panels by use of the CD45^+^ leukocyte ‘backbone’ in combination with the cell count obtained from all samples. Cells were plotted using color density bi-exponential displays to ensure correct identification of negative and positive cell populations.

### PBMC Isolation and Cryopreservation

All steps of peripheral blood mononuclear cells (PBMC) preparation were carried out at room temperature. The content of the blood collecting tubes from the same probands were pooled and mixed 1:1 with pre-warmed 1×PBS. The blood:PBS mixture was carefully layered onto Biocoll (Biochrom) and subsequently centrifuged for density gradient at 1000 g for 20 min, at room temperature (slow acceleration, no brake). The ring-shape interphase (containing PBMCs) was collected with a Pasteur pipette into a new 50 ml tube and diluted up to 50 ml with pre-warmed 1×PBS and centrifuged with 300 g for 10 min, at room temperature, with fast acceleration and with brake. The number of living cells was determined with Trypan blue exclusion as number of living cells/ml of blood. Following a further washing step in 1×PBS (300 g, 10 min, room temperature), cells were resuspended in freezing media containing fetal calf serum (FCS, PAA) with 10 % dimethyl-sulfoxide (DMSO, Sigma-Aldrich) 1 ml aliquots with 5×10^6^ cells/ml were gradually frozen in a freezing container at −80°C for at least 48 hours and transferred into a cryo tank (liquid nitrogen vapor phase).

### T Cell Receptor Sequencing

Genomic DNA was isolated using the QIAamp DNA Mini Kit (Qiagen) according to manufacturer’s instructions for sequencing of T-cell receptors (TCRs). Libraries for TCR-β subunit profiling were prepared utilizing the hsTCRb Kit (Adaptive Biotechnologies) following the manufacturer’s instructions and sequencing was performed on Illumina. Thereafter, ImmunoSEQ-Analyzer 3.0 software was used for sequencing data analysis. Briefly, the most variable complementary-determining region 3 (CDR3), spanning the recombination site of V-D-J recombinations of TCR-β-chains was sequenced, as previously described (*41*). Reads with an average quality score below 20 were removed from the analysis. The remaining reads were processed and further analyzed using IMSEQ as previously performed by Kuchenbecker et al. (*42*). For estimating the diversity of the TCR repertoire, the Shannon Entropy option was used (ShannonWienerIndex_mean value/100).

### Cytokine Assay

The protein levels of IL-6, IL-13, IL-7, IL-8, MIP-1a, MIP-1b, MIP-3a, ITAC and Fractalkine in SAT lysates were measured using the MILLIPLEX® MAP 384-Well High Sensitivity Human Cytokine Magnetic Bead Panel (Millipore Sigma).

### Statistical Analysis

The results are shown as the mean ± SEM. All statistical analysis was performed using GraphPad Prism version 8 (GraphPad Software, San Diego, CA) and R version 3.2.1, available free online at https://www.r-project.org. Unless otherwise notified, no mathematical correction was made for multiple comparisons in order to reduce the error of the second kind. Depending on the distribution of data, Pearson simple coefficient or Spearman rank correlation coefficient were used for correlation analysis. Mann-Whitney *U* test or Student *t* test were applied to estimate differences between groups. Repeated-measures two-way ANOVA, multiple testing with Bonferroni correction and a linear mixed effector model with False discovery rate (FDR) and Bonferroni correction were used to analyze time courses. Significant differences were assumed for P < 0.05 (two-tailed).

## Results

### Obese Probands Have a Decreased Proportion of Effector Memory CD8^+^ T Cells Associated with an Enhanced CD4/8 T Cell Ratio in Peripheral Blood

First, we performed multiparameter flow cytometric analyses to determine whether the composition of leucocyte cell subpopulations is affected in the peripheral blood of obese probands at the starting time point of the study (M0) compared to age and sex-matched healthy lean probands (n=21 vs. 16). For this purpose we employed the so-called “ONE study” flow cytometry panels consisting of six leucocyte profiling 10-color panels, which has been designed and extensively validated in multicenter trials in our lab (*40*). We did not reveal significant differences in the numbers of total leucocytes or major leucocyte subpopulations (Figure 2A). Subset analyses, however, showed an increased percentage of activated monocytes (CD14^+^CD16^high^) in obese subjects (*P* = 0.020, data not shown), suggesting a proinflammatory state, which is consistent with previous literature (*7-9*). However, we observed an increase of the naïve compartment of CD4^+^ and CD8^+^ T cells within the circulating T cells in the obese cohort whereas percentages of central memory, effector memory and terminally differentiated T cells were not significantly different between lean and obese probands (Figure 2B). Also, absolute counts of naïve CD4^+^ T cells were increased (Figure 2C) whereas effector memory CD8^+^ T cells (Figure 2C) were reduced in obese probands. The analysis of B cell subtypes showed that obese subjects have higher numbers of naïve B cells (Figure 2D). These results are partly in line with recently published observations (*43*) demonstrating a selective increase of peripheral blood CD4^+^ T cells skewed toward an anti-inflammatory phenotype (CD4^+^ naïve, natural CD4^+^CD25^+^FoxP3^+^Treg, and Th2 T cells) in morbidly obese human subjects. Taken together, our data demonstrate that obesity is associated with a shift toward naïve T cells in both CD4^+^ and CD8^+^ compartments in peripheral blood.

**Figure 2.**
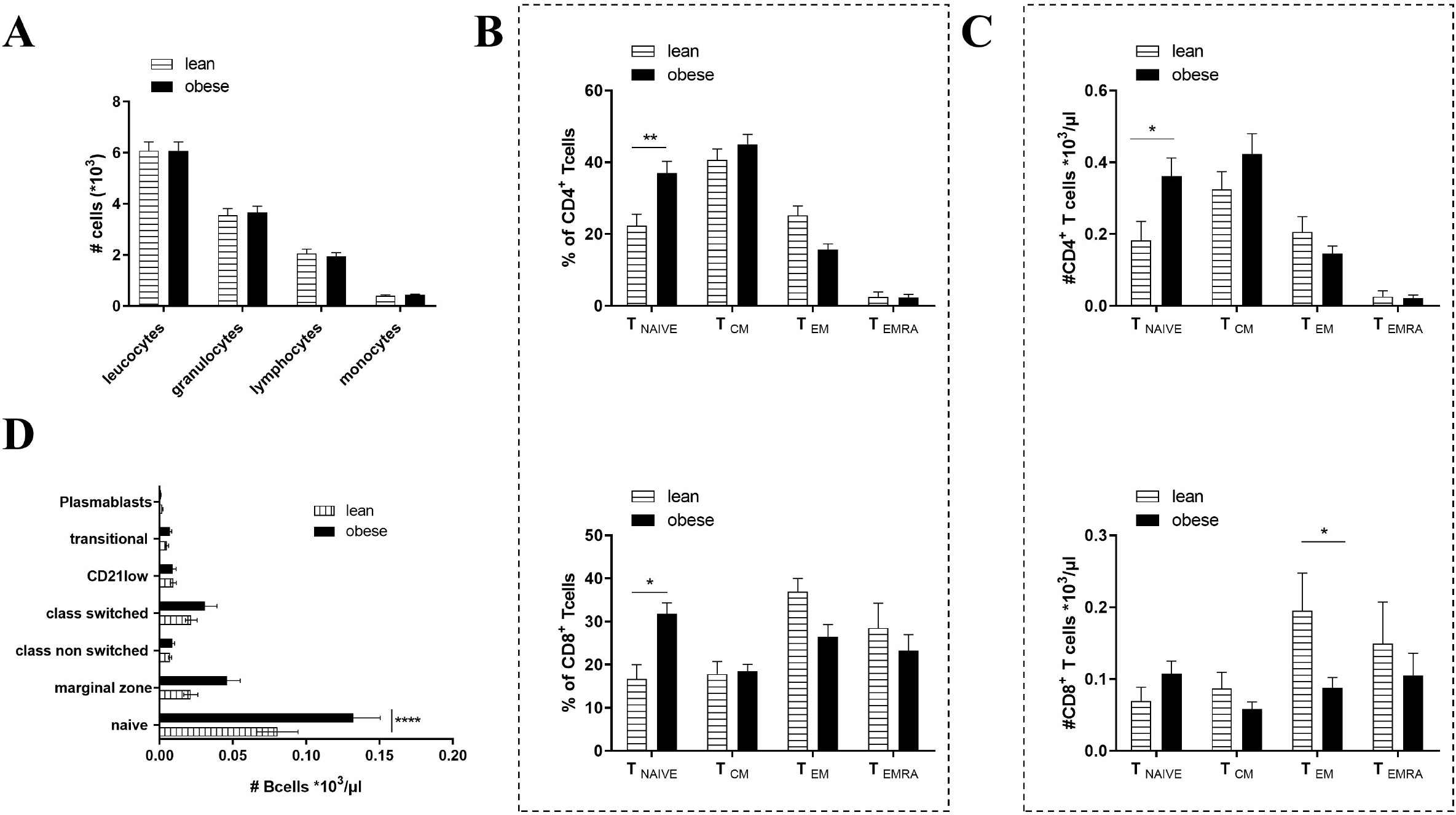
Altered immune cell composition in peripheral blood in overweight/obese vs. lean probands. PBMCs isolated from venous blood of overweight/obese (n=21) and lean (n=16) age and sex-matched probands were analyzed with multiparameter flow cytometry at time point 0 (M0). **(A)** No differences among absolute major leukocyte subset counts were observed. **(B, C)**. Proportions of CD4^+^ and CD8^+^ T cells and absolute cell counts of CD4^+^ and CD8^+^ naïve (T_naïve_), central memory (T_CM_), effector memory (T_EM_) and terminally differentiated effector memory T cells (T_EMRA_) determined from flow cytometric data. An increase in T_naïve_ but decrease in T_EM_ was observed for both percentages of CD4^+^ and CD8^+^ T cells and absolute cell counts in overweight/obese compared to lean probands. **(D)** Absolute cell counts of B cells showed an increase of naïve B cells in overweight/obese compared to lean probands. For all graphs data points were tested with repeated measurement 2way ANOVA followed by SIDAK correction. *P*-values below 0.05 are indicated by * and defined to be significant.

### Alterations of Systemic Leucocyte Subpopulations Correlate with Metabolic Measures in Obesity

To further assess the association of metabolic parameters with the altered systemic immune cell composition in obesity, we examined the correlations between BMI, insulin sensitivity index (ISI_Clamp_), waist circumference and leucocyte cell numbers of lean and overweight/obese probands of the sub-study C. A significant correlation was found between the BMI and total leucocyte numbers (*r* = 0.367, *P*= 0.035, Figure 3A). Moreover, BMI correlated positively with the number of activated CD14^+^CD16^high^ monocytes (*r* = 0.459, *P* = 0.007, Figure 3B). Additionally, we found positive correlations for BMI with naïve and central memory CD4^+^ T-cell numbers (*r* = 0.401, *P* = 0.046; *r*= 0.682, *P* < 0.001, Figures 3C, D) but not with effector memory or terminally differentiated effector memory CD4^+^ T cell numbers (Supplementary Table 1). Similarly, a significant correlation between BMI and naïve CD8^+^ T-cell numbers (*r* = 0.483, *P* = 0.014, Figure 3E) but not with other CD8^+^ T-cell subsets (Figure 3F, Supplementary Table 1) was found. Additionally, absolute numbers of lymphocytes (*r* = 0.494, *P* = 0.027), T cells (*r* = 0.545, *P* = 0.013), CD4^+^ (*r* = 0.475, *P* = 0.034) and CD8^+^ T cells (*r* = 0.452, *P* = 0.045), and TCRαβ cells (*r* = 0.564, *P* = 0.009) correlated positively with waist circumference (Supplementary Table 1). Insulin and glucose levels were determined in the overweight/obese control and CR group of the sub-Study C. The ISI_Clamp_ was then calculated for all obese probands as a measure of insulin-mediated glucose uptake. Only total leucocyte (*r* = −0.482, *P* = 0.031) and monocyte (*r* = −0.519, *P* = 0.019) cell counts correlated negatively (and thus reciprocally with BMI) with ISI_Clamp_ (Figure 3G, H). While, the percentage of B cells correlated positively with ISI_Clamp_ (*r* = 0.498, *P*= 0.022, Figure 3I), in the T cell compartment, a significant correlation could only be found for the percentage of naïve CD8^+^ T cells (*r* = 0.561, *P*= 0.008, Figure 3J, Supplementary Table 2). Using chemokine receptor panels, we further analyzed the distribution of functional T cell subsets. Recently, a role of Th17 and Th22 cells in obesity-mediated IR has been described (*21*). Th17-derived Th1 cells have been termed non-classic Th1 cells and can be identified based on their constant CCR6 expression. Here, we found a significant correlation of non-classic Th1 cells with ISI_Clamp_ (*r* = 0.662, *P*= 0.002, Figure 3K). These results are not consistent with published reports showing that adipose-associated Th1 cells promote obesity-associated inflammation (*44, 45*). To determine whether the shift to a more naïve T-cell composition in peripheral blood of obese probands is due to migration of the effector memory T cells to the adipose tissues, we next studied intra-SAT cell composition.

**Figure 3.**
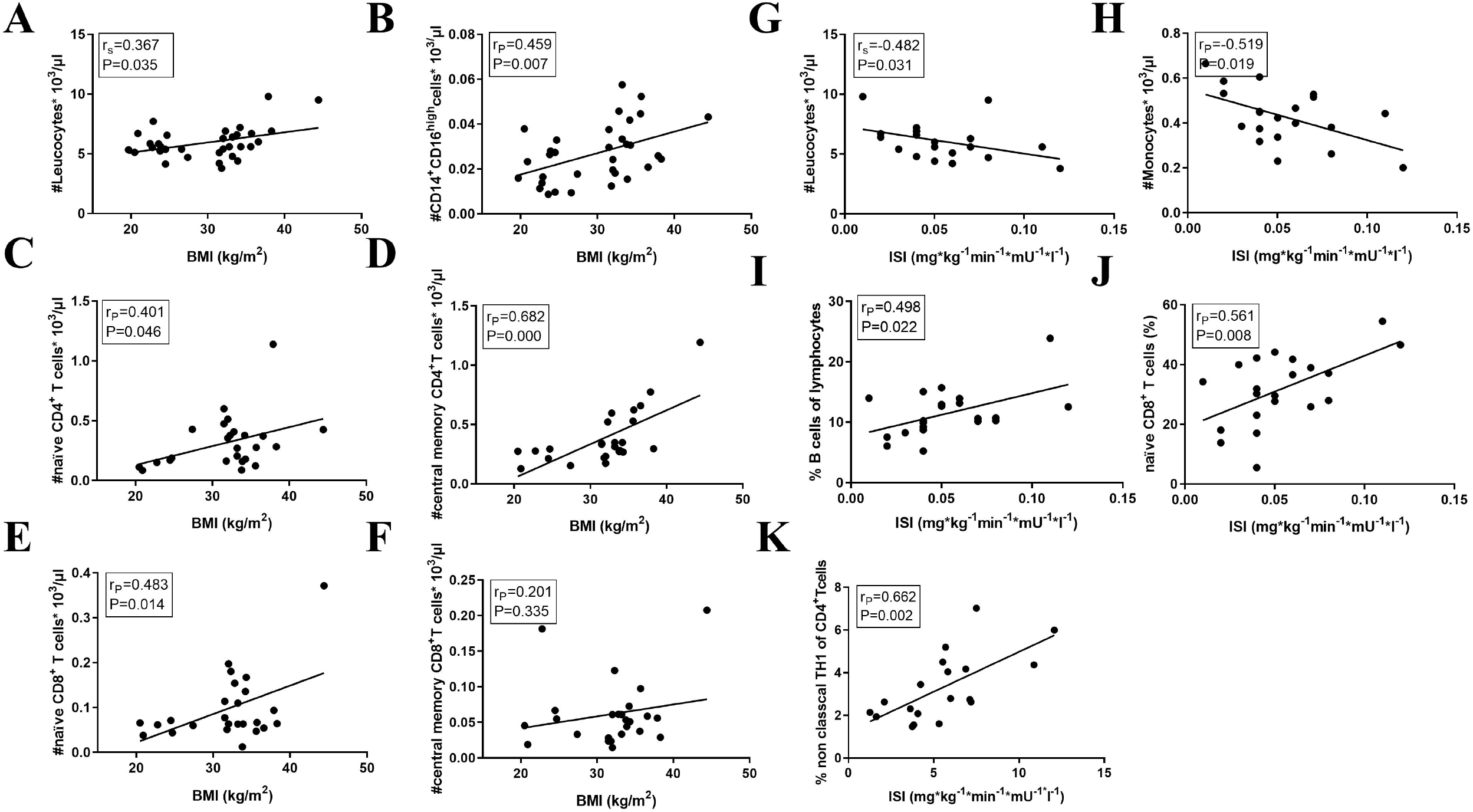
Altered composition of immune cell subsets in obesity is correlated with the BMI. PBMCs isolated from venous blood of overweight/obese (n=21) and lean (n=16) age and sex-matched probands were analyzed with multiparameter flow cytometry at time point 0 (M0) and resulting immune cell parameters were correlated with metabolic measures. **(A-K)** Positive correlations between peripheral blood leukocyte counts **(A)**, activated monocytes (CD14^+^CD16^high^) **(B)**, naïve CD4^+^ **(C)**, early central memory CD4^+^ **(D)**, naïve CD8^+^ **(E)**, early central memory CD8^+^ **(F)** T-cell subsets and the BMI. Negative correlations between leucocyte and monocyte cell counts with ISI_Clamp_ **(G, H)**. Positive correlation between the percentage of B cells **(I)**, naïve CD8^+^ T cells **(J)** and non-classical Th1 CD4^+^ T cells **(K)** and ISI_Clamp_. Black dots represent study probands (obese n=21, lean n=16). Spearman’s and Pearson’s correlation analysis was conducted.

### Intra-SAT Accumulation of Effector Memory T Cells Correlates with ISI in Obesity

To ascertain the relationship between T cell profiles and metabolic measures during obesity, we investigated the correlations between BMI, waist circumference and systemic IR and leucocyte cell numbers in SAT of overweight/obese probands of the sub-study C. To our knowledge, this is the first reported broad characterization of immune cell subsets in human SAT using a highly standardized assay. We observed an increased percentage of granulocytes in SAT compared to paired blood samples, but decreased lymphocyte counts (Figure 4A). Interestingly, within both the CD4^+^ and the CD8^+^ T-cell compartment, increased percentages of effector memory T cells were found in SAT, whereas naïve and central memory T-cell percentages were decreased and no differences were found in terminally differentiated effector memory T cells (TEMRAs) (Figures 4B-D). Interestingly, naïve and effector memory CD8^+^ T cells, but no other memory and effector T-cell subsets correlated with the ISI_Clamp_ (*r* = 0.485, *P* = 0.026; *r* = −0.738, *P* < 0.001, Figures 4E-F). Although the frequency of central memory CD4^+^ and CD8^+^ T cells was lower in SAT compared to paired blood samples, they correlated positively with the BMI (*r* = 0.548, *P* = 0.010; *r* = 0.482, *P* = 0.027, respectively) whereas TEMRA cell number was inversely correlated with BMI (*r*= −0.566, *P* = 0.008, Supplementary Table 3, respectively). No significant correlations with metabolic measures (BMI, waist circumference, ISI_Clamp_) were found between other leucocyte subsets (Supplementary Table 3). Inconsistent with previous results (*24, 46*), we could hardly detect any Th22-like or Th17-like cells, although they were detectable in peripheral blood (data not shown). However, all other immune cell subsets were consistently detectable. Overall, these results suggest that the accumulation of effector memory T cells in human SAT is associated with IR. The drop in effector memory T cell counts in the blood of the obese probands might result from migration into the inflamed SAT.

**Figure 4.**
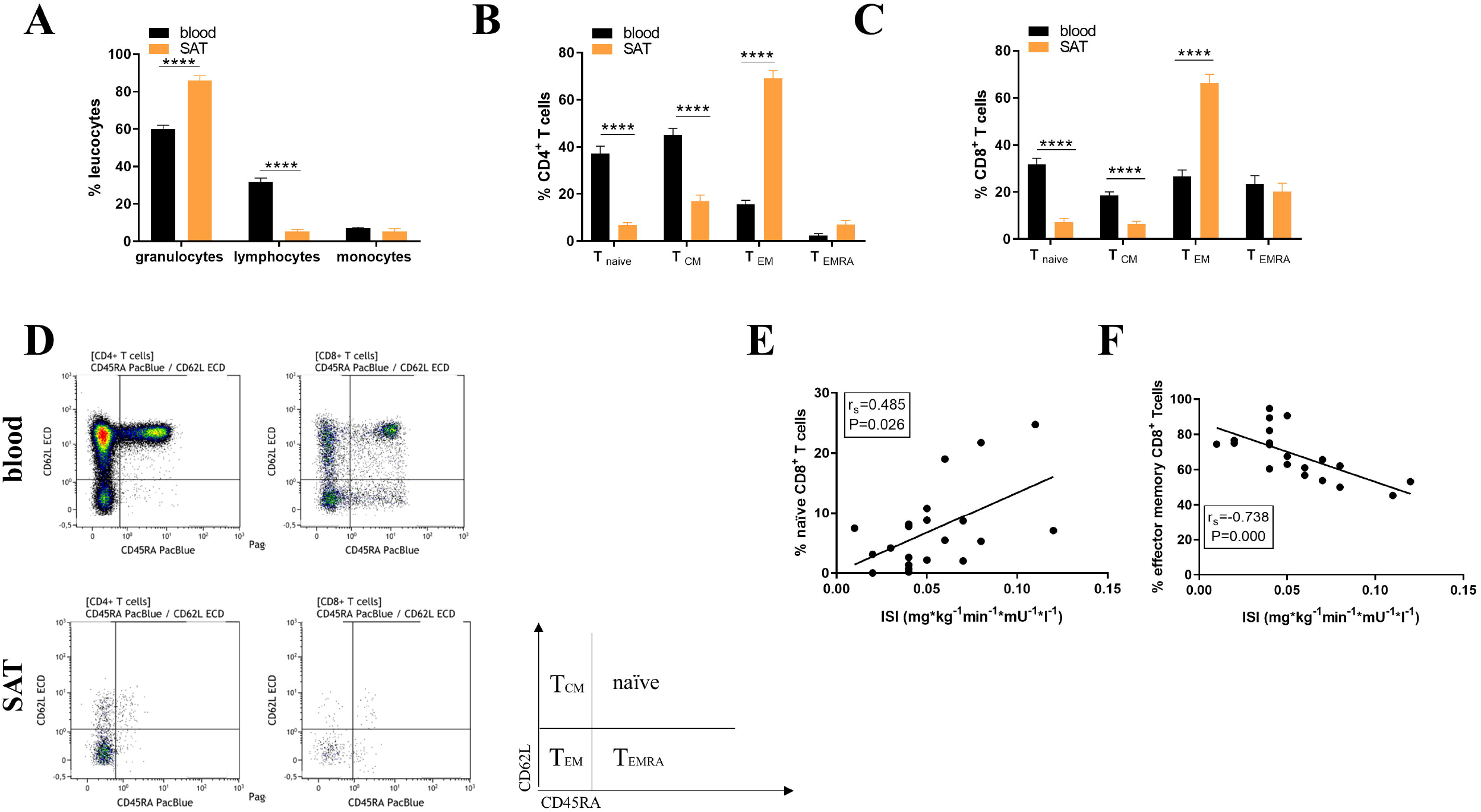
Influence of obesity on different T cell memory subsets in human SAT. SAT immune cells from overweight/obese age and sex-matched probands (n=21) were isolated and analyzed with multiparameter flow cytometry at time point 0 (M0). Total cell counts and percentages were compared between peripheral blood and SAT. **(A-C)** Leucocyte and T cell subpopulations in blood and SAT of obese probands. **(D)** Representative dot plots of CD4^+^ and CD8^+^ T cell subsets in blood and SAT of obese probands. **(E, F)** Correlations between CD8^+^ T cell subsets with the ISI_Clamp_. *** P < 0.001 **** P < 0.0001 Spearman’s correlation analysis was conducted.

### Characterization of the SAT TCR-β Repertoires of Obese Probands Revealed Preserved TCR Diversity

T cells can be identified via their unique TCR sequence. Given the involvement of the adaptive immune system and strong correlations of effector memory T cell subsets with metabolic parameters, we analyzed the TCR repertoire of SAT and blood derived T cells to address the question whether unspecific bystander expansion or an (auto)-antigen-driven process is the main driver of effector memory T cell accumulation in obese SAT. We compared clonal relations between SAT and blood derived T cells of 20 obese probands (sub-study B) at M0. TCR-β chain repertoire analysis of adipose T cells by next generation sequencing revealed a polyclonal repertoire with overlapping TCRs with peripheral blood T cells, suggesting that there was no strong skewing towards particular T cell clones or a unique SAT repertoire. Applying the Shannon Entropy algorithm (*47*) and normalizing the number of unique T cell clonotypes by the total number of sequences per sample, the diversity of the TCR repertoire was assessed using the total number of unique TCR-β clonotypes in blood and SAT samples and revealed a very similar degree of diversity with values close to 1, as shown in Figure 5A. The frequencies of the top100 clones in SAT and blood are shown in Figure 5B.

**Figure 5.**
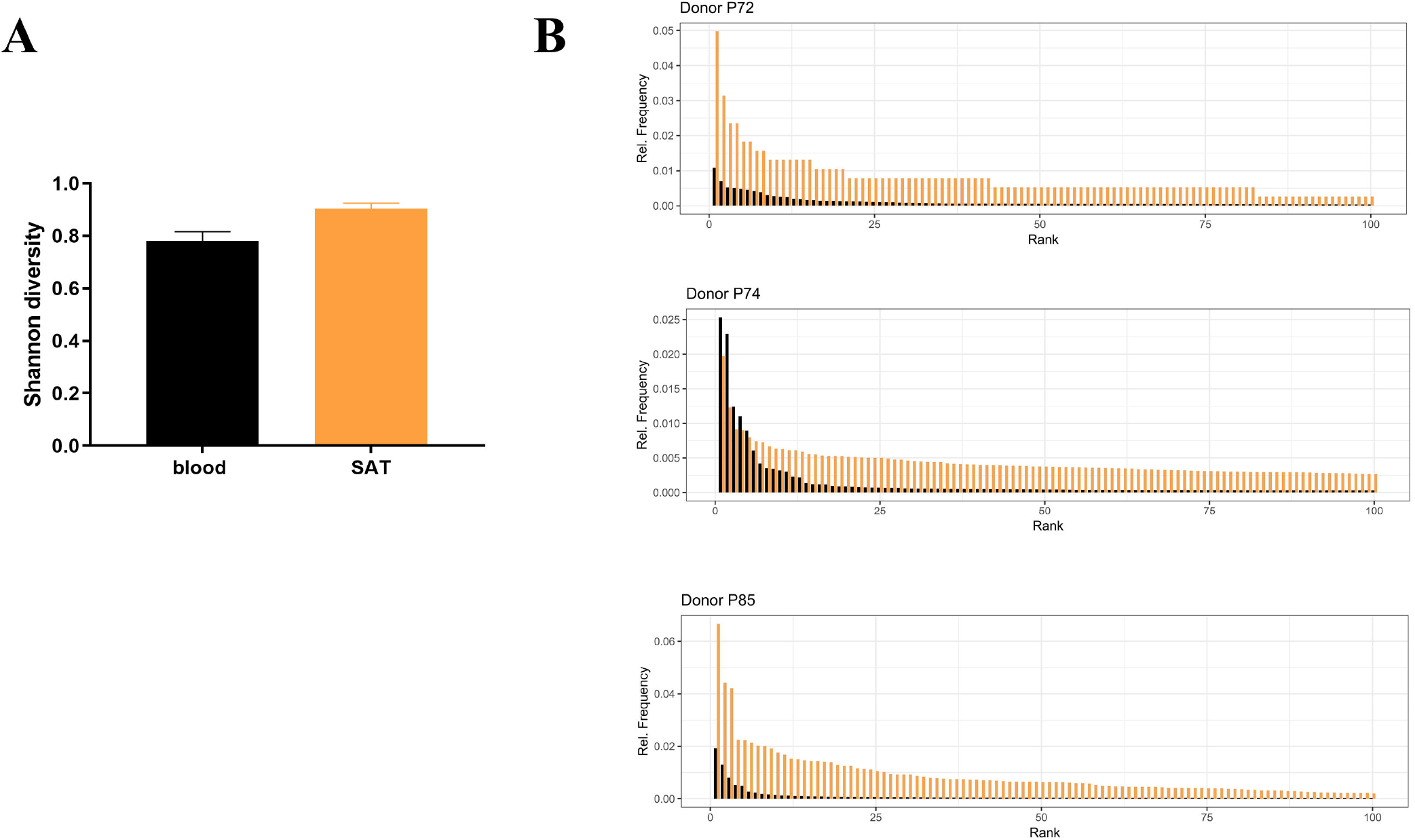
TCR clonal expansion and diversity of TCR repertoire in SAT and blood of obese patients. **(A)** The diversity of the TCR repertoire from peripheral blood and SAT from obese patients (n=20) from sub-study B. The mean and standard deviation for the entropy value when sampling the same number of sequences from the blood repertoire as obtained from the SAT repertoire. The difference between the observed entropy in the SAT samples and the mean entropy from sampled repertoires is provided in the last column. **(B)** Frequencies among productive rearrangements of the top100 clones of SAT (orange) and Blood (black) of 3 overweight/obese probands. ** P <0.01. Comparison between blood and SAT samples were tested by Wilcoxon signed-rank test.

### Short-Term CR Does Not Reverse the Shift of Effector Memory T Cells from Blood to SAT and the Local Release of Inflammatory Cytokines Despite Positive Metabolic Effects

To assess the effects of CR on immunological alterations, we investigated the kinetics of leucocyte subsets in blood and SAT of obese probands prior to weight loss and then 3 and 4 months after compared to a control group that maintained a stable weight (sub-study C). Interestingly, we did not find any significant decrease in the total number of leucocytes after correction for multiple testing with Bonferroni or FDR although they were highly correlated with BMI (Figure 6A). The linear mixed model testing for group x time interactions for total numbers and percentages of leucocyte subsets revealed no significant group x time interactions at time points M3 and M4 in SAT or peripheral blood indicating no significant difference in leucocyte subset composition between the groups after the weight reduction and weight maintenance phase in the intervention group in comparison with the control group. Most notably, the percentage of all T cell subsets, including naïve, central memory, effector memory and terminally differentiated effector memory T cells, remained completely unaffected (Supplementary Table 4). We next studied the impact of CR on intra-SAT cytokine levels with a multiplex assay in 22 obese probands (sub-study A), as the association between obesity and increased release of pro-inflammatory cytokines is well known. MIP-1a correlated positively with the BMI (*r* = 0.588, *P* = 0.018, Figure 6K), while MIP-3a correlated negatively with ISI_Clamp_ (r = −0.433, *P* = 0.044). Both cytokines are inducible C-C family chemokines that play a pivotal role in the recruitment of monocytes, macrophages (MIP-1a) and lymphocytes (MIP-3a) (*48, 49*) (Figure 6L and Supplementary Table 5). Additionally, HOMA correlated positively with MIP-1a (r = 0.584, *P* = 0.009), MIP-1b (r = 0.555, *P* = 0.011) and MIP-3a (r = 0.524, *P* = 0.018) (data not shown). To our surprise, intra-SAT levels of IL-6, IL-13, IL-7, IL-8, MIP-1a, MIP-1b, MIP-3a, ITAC and Fractalkine were not significantly influenced by CR-induced weight loss, as shown in Figure 6B-J. Taken together, our data show that the immune cell phenotype in blood and SAT is not substantially altered after CR despite metabolic improvements.

**Figure 6.**
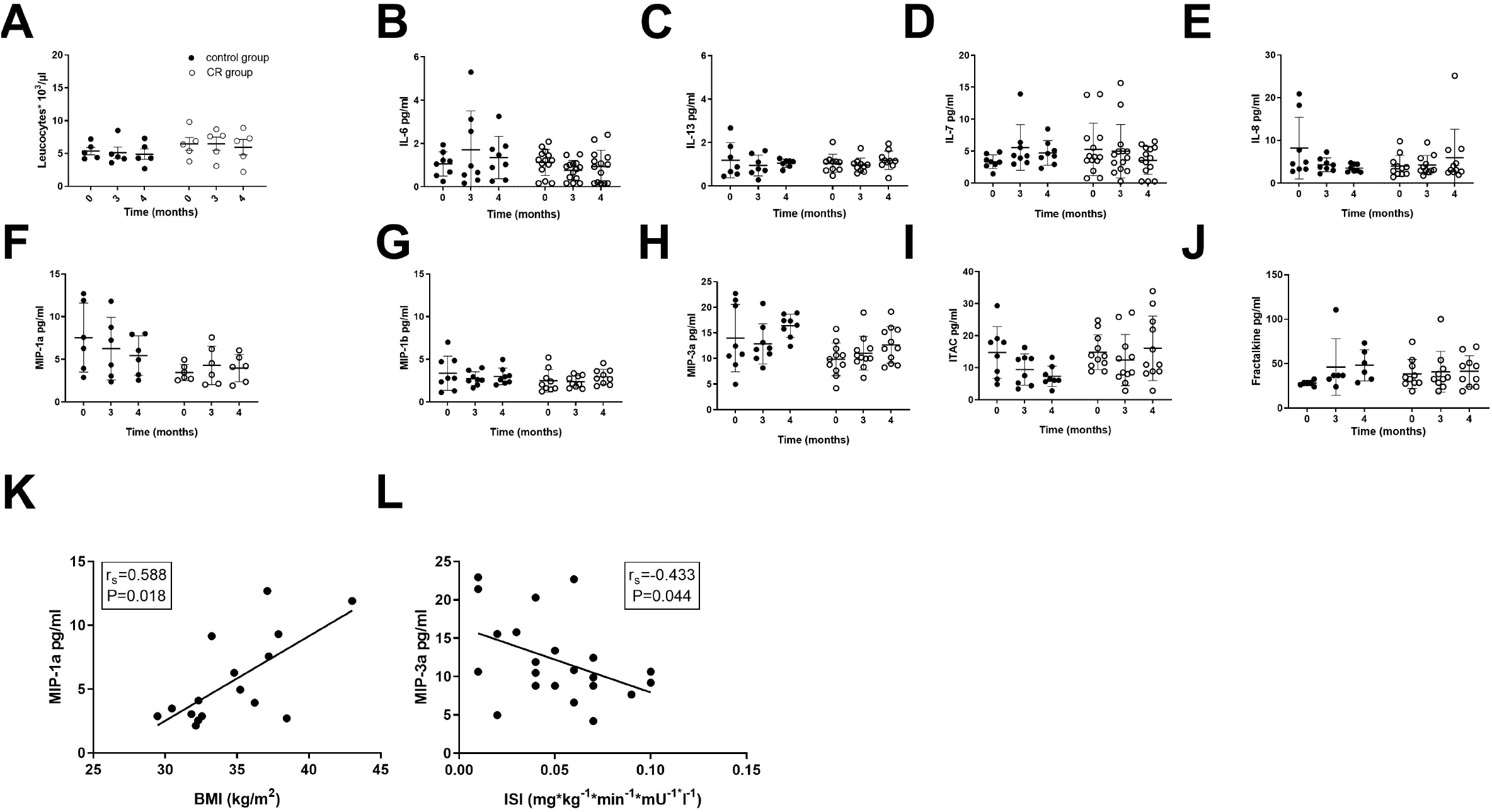
Leucocyte numbers and cytokine levels in SAT are not altered after CR. **(A)** Systemic leucocyte cell numbers at time points 0 (M0, starting point), 3 (M3) and 4 (M4) in overweight/obese probands (n=22) of the control (black dots) and CR group (white dots). **(B-J)** SAT levels of IL-6, IL-13, IL-7, IL-8, MIP-1a, MIP-1b, MIP-3a, ITAC and Fractalkine in probands of the control and CR group. **(K)** Correlation of BMI with MIP-1a levels in SAT of probands of the control and CR group. **(L)** Correlation of ISI_Clamp_ with MIP-3a levels in SAT of probands of the control and CR group. Spearman’s correlation analysis was conducted. *P*-values below 0.05 are indicated by * and defined to be significant.

## Discussion

This study performs, for the first time, a comprehensive immune monitoring of human blood and SAT tissue using flow cytometry in a standardized assay, which includes six leucocyte profiling panels and which yields acceptable variability tested at multiple international sites (*40*), allowing unmanipulated analysis of changes in absolute cell numbers of leukocyte subsets in SAT of obese individuals. Although impressive findings have been demonstrated in rodent studies, the immune cell composition in human WAT is unclear and there are no studies providing standardized immune monitoring of leucocyte subsets in human WAT before and after weight loss induced by CR. To our knowledge, there are only few studies investigating various T cell subsets in human WAT using flow cytometry. First, increased prevalence of Th17 and Th22 cells has been shown in SAT of metabolically abnormal versus metabolically healthy obese subjects (*24*). Second, a specific accumulation of IL-22 and IL-17 producing CD4^+^ T cells was found in SAT of type 2 diabetic obese patients (*46*). Furthermore, an increased frequency of Th17 cells was demonstrated in SAT of overweight versus lean women (*50*). Flow cytometry revealed significant correlations between waist circumference and levels of activation marker expression on CD4^+^ and CD8^+^ T cells in SAT (*51*). Only recently, the association of CD4^+^ T cells in visceral WAT with their counterparts in SAT was investigated in lean and obese men (*52*).

We confirm prior studies demonstrating various T cells are present in human adipose tissues. Importantly, our results demonstrate that subpopulations of T cells are associated with both body weight and systemic IR. In obese women, pro-inflammatory effector memory CD8^+^ T cells were markedly increased in SAT relative to peripheral blood, which was associated with a shift to enhanced levels of naïve T cells in the circulation. Numbers of both naïve and effector memory T cells showed robust bidirectional relationships to BMI. Importantly, IR correlates positively with effector memory CD8^+^ T-cell counts and negatively with naïve CD8^+^ T cells counts in SAT. Due to their specific chemokine, adhesion, and homing receptor profile, effector memory T cells can enter any given inflamed site, while naïve and central memory T cells circulate in the blood and the lymphatic system. Moreover, effector memory T cells can be activated easier by bystander activation and do not depend on TCR signals. The release of proinflammatory cytokines, such as TNFα and IFNγ can trigger inflammation with accumulation and activation of other leucocyte subsets, such as myeloid cells further amplifying inflammation in adipose tissues. In fact, SAT expression of MIP-1a (as well as MIP-3b and MIP-3a), a chemokine recruiting monocytes and macrophages to peripheral tissues, was highly associated with BMI. Altogether, the relative dominance of antigen-experienced effector memory versus naïve T cells in obese SAT may account in part for the metabolic differences described in obese humans. The present findings contribute to our understanding of the link between inflammatory cells and the development of IR. Previously, the majority of studies related to inflammation and IR focused on innate immunity, but recent studies in mice demonstrated an important role of CD4^+^ and CD8^+^ T cells in the development of IR (*18, 53, 54*). They have shown that cytotoxic CD8^+^ T cells and Treg cells play opposing roles in IR and WAT inflammation. In line with these findings, we previously observed that effector memory CD8^+^ T cells correlate significantly with glucose tolerance, measured via intraperitoneal glucose tolerance tests in high-fat diet fed mice (*55*). In conclusion, the present study supports the hypothesis that proinflammatory immune action in SAT plays an important role in determining IR.

While there is evidence that adaptive immunity is involved in the development of IR in mice, it is unclear if the accumulation of B and T cells in WAT is based on their antigen-specific activation or functions as a result of general inflammation. Importantly, some of the above mentioned studies showed severely biased T-cell repertoires in WAT (*53, 56*), suggesting intra-WAT antigen-specific responses of T cells. Hypertrophic adipocytes can become necrotic and release cellular components and secrete molecules, which could serve as potential (auto-)antigens. Stress proteins are also discussed as T cell targets. In contrast to these mice studies, no significant changes to the TCR repertoire were observed in a recently published human study using GeneScan analysis of Vβ-Jβ arrangements to compare lean versus non-diabetic obese subjects (*43*). In line with these findings, we found a polyclonal TCR-β repertoire in SAT of obese probands using next generation sequencing.

Obesity has become one of the most serious health problems globally. Whereas it is not difficult to lose weight, only a small minority is able to maintain the reduced weight long-term. Surprisingly, we were unable to detect significant alterations in frequencies of leucocyte subsets investigated via flow cytometry or secretion of cytokines investigated via immunoassay after short-term CR induced weight loss. Consistently, it has been previously reported that mice with obesity experience displayed much faster body weight regain, while this obesity memory had a long-lasting effect and was mainly mediated by CD4^+^ T cells from previously obese mice (*57*). Additionally, previous mouse studies reported that weight fluctuation enhances inflammation due to persistent microbiome alterations in obese mice (*58*). These results suggest that T cells have a vital role in establishing obesity memory and might explain why short-term CR of obese probands often results in unavoidable weight regain.

Our study is limited by the relatively small number of obese subjects willing to undergo CR intervention as well as SAT biopsies and the inability to perform extensive flow cytometry on every tissue sample at all three intervention time points of the sub-study C. However, as we investigated a very specific sample, the relative homogeneity of our study participants (obese postmenopausal women) could strengthen the present study (small range in BMI and age is important due to age-related changes in immune function). Nevertheless, future studies are needed to extend the findings to a broader population. Moreover, as is common in observational human studies, it is difficult to prove causality. However, considering the present findings and those from prior weight gain-loss obese mice studies (*57*), in which CD4^+^ T cells were shown to mediate obesity memory and promote weight regain, our results are highly suggestive of a similar relationship in humans. As the gut microbiota seems to be involved in the induction of obesity and interacts with the gut immune system (*59*), regulating the formation of effector memory T-cell subsets, studies investigating the modulation of the gut microbiome through diet, pre- and probiotics, antibiotics, surgery, and fecal transplantation are warranted. The modulation of the gut microbiome through CR-induced weight loss was investigated in another sub-study in our group (data submitted).

In summary, the present findings extend previous studies in mice and add to the limited reports in humans, implicating a role of T cells in obesity-related IR and T2D and providing evidence for a link between inflammation and IR in humans. Moreover, we provide new insight into the impact of CR on the inflammatory state in SAT. Collectively, these data indicate that the inflammatory signature in SAT of obese probands could not be reshaped by CR. Further investigation into specific T lymphocyte subtypes and antigens involved in the mechanism of CR and weight regain, may help to develop more efficient anti-obesity strategies.

## Data Availability

The raw data supporting the conclusions of this manuscript will be made available by the authors, without undue reservation, to any qualified researcher

## Conflict of Interest

The authors declare that the research was conducted in the absence of any commercial or financial relationships that could be construed as a potential conflict of interest.

## Author Contributions

JS-K, H-DV and JS designed and planned the study. JK performed experiments supervised by JS-K. JS-K performed experiments, analysed and interpreted data. JS-K composed figures and JS-K, H-DV and JS wrote the manuscript. KM, RJvonS, A-MD and LS provided patient samples and data, MS provided help with flow cytometry analysis, LK and KJ performed bioinformatic analyses, UK helped with the sequencing devices and all authors approved the final version of the manuscript.

## Funding

JS was supported by the DZHK (German Center for Cardiovascular Research), JS, KM and J-SK were supported by a clinical research group of the DFG (KFO218 and KFO192). This study was supported by grants from the Clinical Research Unit of the Berlin Institute of Health (BIH), the “BCRT-grant” by the German Federal Ministry of Education and Research and the Einstein Foundation. JSK and RJvonS were supported by the Charité Clinical Scientist Program.

## Acknowledgments

We thank Francesca Liersch, Sarina Richter and Christiane Gras for excellent assistance with experimental procedures. We acknowledge the assistance of the BCRT Flow Cytometry Core Facility, Dr. D. Kunkel and Dr. Sarah Meier. We thank Catherine Steer for carefully reading the manuscript.

**The raw data supporting the conclusions of this manuscript will be made available by the authors, without undue reservation, to any qualified researcher**.

## Notes

### Competing Interest Statement

The authors have declared no competing interest.

### Clinical Trial

NCT01105143

### Author Declarations

All relevant ethical guidelines have been followed and any necessary IRB and/or ethics committee approvals have been obtained.

Any clinical trials involved have been registered with an ICMJE-approved registry such as ClinicalTrials.gov and the trial ID is included in the manuscript.

